# Clinical signatures of genetic epilepsy precede diagnosis in electronic medical records of 32,000 individuals

**DOI:** 10.1101/2022.12.08.22283226

**Authors:** Peter D. Galer, Shridhar Parthasarathy, Julie Xian, Jillian L. McKee, Sarah M. Ruggiero, Shiva Ganesan, David Lewis-Smith, Michael C. Kaufman, Stacey R. Cohen, Scott Haag, Alexander K. Gonzalez, Olivia Wilmarth, Colin A. Ellis, Brian Litt, Ingo Helbig

**Affiliations:** Division of Neurology, Children’s Hospital of Philadelphia, Philadelphia, PA, USA; Department of Biomedical and Health Informatics (DBHi), Children’s Hospital of Philadelphia, Philadelphia, PA, USA; The Epilepsy NeuroGenetics Initiative (ENGIN), Children’s Hospital of Philadelphia, Philadelphia, PA, USA; University of Pennsylvania, Center for Neuroengineering and Therapeutics, Philadelphia, PA, USA; Department of Neurology, University of Pennsylvania Perelman School of Medicine, Philadelphia, PA, USA; Translational and Clinical Research Institute, Newcastle University, UK; Newcastle Upon Tyne Hospitals NHS Foundation Trust, Newcastle-upon-Tyne, UK

**Keywords:** precision medicine, epilepsy, genetics, developmental and epileptic encephalopathy, Human Phenotype Ontology

## Abstract

An early genetic diagnosis can guide the time-sensitive treatment and care of individuals with genetic epilepsies. However, identification of a genetic cause often occurs long after onset of these disorders. Here, we aimed to identify early clinical features suggestive of genetic diagnoses in individuals with epilepsy by systematic large-scale analysis of clinical information from full-text patient notes in the electronic medical records (EMR).

From the EMR of 32,112 individuals with childhood epilepsy, we retrieved 4,572,783 clinical notes spanning 203,369 total patient-years. A subcohort of 1,925 individuals had a known or presumed genetic epilepsy with 738 genetic diagnoses spanning 271 genes. We employed a customized natural language processing (NLP) pipeline to extract 89 million time-stamped standardized clinical annotations from free text of the retrieved clinical notes. Our analyses identified 47,641 clinical associations with a genetic cause at distinct ages prior to diagnosis. Notable among these associations were: *SCN1A* with status epilepticus between 9 and 12 months of age (*P*<0.0001, 95% CI=8.10-133); *STXBP1* with muscular hypotonia between 6 and 9 months (*P*=3.4×10^−4^, 95% CI=3.08-102); *SCN2A* with autism between 1.5 and 1.75 years (*P*<0.0001, 95% CI=11.1-Inf); *DEPDC5* with focal-onset seizure between 5.75 and 6 years (*P*<0.0001, 95% CI=12.8-Inf); and *IQSEC2* with myoclonic seizure between 2.75 and 3 years (*P*=2.5×10^−4^, 95% CI=11.3-1.15×10^4^). We also identified associations between clinical terms and gene groups. Variants in ion channel gating mechanisms were associated with myoclonus between 3 and 6 months of age (*P*<0.0001, 95% CI=5.23-24.2), and variants in calcium channel genes were associated with neurodevelopmental delay between 1.75 and 2 years (*P*<0.0001, 95% CI=4.8-Inf). Cumulative longitudinal analysis revealed further associations, including *KCNT1* with migrating focal seizures from at 0 to 1.75 years (*P*<0.0001, 95% CI=96.8-4.50×10^15^). A neurodevelopmental abnormality presenting between 6 and 9 months of age was strongly associated with an individual having any genetic diagnosis (*P*<0.0001, 95% CI=3.55-7.42). The earliest features associated with genetic diagnosis occurred a median of 3.6 years prior to the median age of diagnosis. Latency to diagnosis was greater in older individuals (*P*<0.0001) and those who initially underwent less comprehensive genetic testing (*P*=5.5×10^−3^, 95% CI=1.23-3.35).

In summary, we identified key clinical features that precede genetic diagnosis, leveraging EMR data at scale from a large cohort of individuals with genetic epilepsies. Our findings demonstrate that automated EMR analysis may assist clinical decision making, leading to earlier diagnosis and more precise prognostication and treatment of genetic epilepsies in the precision medicine era.

## Introduction

Childhood epilepsies have a strong genetic basis with more than 100 monogenic etiologies identified as of 2022.^1,2^ Therapeutic advances in child neurology are increasingly tailored towards genetic causes. Thus, an early genetic diagnosis can be critical to inform treatment strategies, contributing to significant treatment change in up to 80% of patients and a decrease in hospitalizations.^3^ However, while some genetic epilepsies have specific features that lend themselves to early recognition and diagnosis, genetic testing is frequently protracted and only performed later in the course of the disease. For example, although clinical features of *SCN1A*-related epilepsy disorders such as Dravet syndrome are typically apparent between 6-9 months, the median age of genetic diagnosis is 4.2 years.^2^ Delays in diagnosis are even greater in genetic epilepsies where clinical features are initially non-specific and compatible with a wide range of genetic causes, such as in Infantile Spasms and West syndrome.^4-6^ In these conditions, a genetic diagnosis is rarely made in the first year of life. Given that many current and future precision medicine strategies ideally will be applied as early as possible during the disease course to affect the underlying pathophysiology, novel strategies are needed for timely patient identification. The prevalence and depth of data within electronic medical records (EMR) may make such advancements in precision medicine possible.

EMR have emerged as a standard in medical documentation. They are used in >85% of hospitals in the United States and are similarly commonplace in the UK, Ireland, Australia, and many other countries.^7-12^ While many EMR applications are primarily built for operational and billing purposes, there is an increasing use of secondary EMR data for research and quality improvement. For example, learning health system frameworks such as Epilepsy Learning Health System (ELHS) championed by the Epilepsy Foundation,^13^ the Pediatric Epilepsy Learning Health System (PELHS),^14^ and the National Clinical Programme for Epilepsy (NCPE)^10,15,16^ in Ireland employ systematically ascertained clinical data to improve ongoing care. We previously demonstrated that many genetic epilepsies could have specific phenotypic signatures in the EMR that may allow for improved prognostication and understanding of treatment responses.^17-19^ Here, we assess whether information captured in the EMR prior to genetic testing can provide clues for the later genetic diagnosis in individuals with epilepsy. We find that a wide range of genetic epilepsies present with prominent clinical features prior to genetic testing and diagnosis. Furthermore, we find that this information can be easily retrieved from the EMR, allowing for automated risk stratification and dedicated genetic testing strategies in children presenting with seizures and neurodevelopmental disorders.

## Materials and methods

### Participant recruitment

All clinical data were derived from Children’s Hospital of Philadelphia (CHOP) Care Network, consisting of a main hospital and 50 satellite clinics. Individuals had at least one EMR within the CHOP Care Network and originated from two cohorts: the Pediatric Epilepsy Learning Health System (PELHS) cohort and Epilepsy Genetics Research Project (EGRP). PELHS is a deidentified cohort consisting of individuals seen within the CHOP Care Network since 2010 who have received a diagnosis before the age of 18 years of any ICD-10/9 epilepsy code, including: ICD9:345.x, ICD9:779.0, ICD10:G40.x, ICD10:R56.x, or ICD10:P90.x. EGRP has been enrolling individuals with a known or presumed genetic epilepsy, collecting both clinical and genetic data since 2014. All genetic diagnoses were assessed in a clinical and research setting and, if necessary, reclassified according to the criteria of the American College of Medical Genetics and Genomics (ACMG). To prevent duplication, all individuals within EGRP were removed from the PELHS cohort by searching for duplicated database identifiers.

### Electronic medical record collection

All encounters are documented in the same integrated EMR system (EPIC, Verona, WI) accessible through the Clarity data warehouse. We retrieved every EMR from the CHOP Care Network from all individuals in both cohorts until March 10, 2022. Records were time-stamped with the age of the individual at the time of the visit. Nearly all medical records outside of our healthcare network were not collected and consequently could not be analyzed. A small number of external records were available in the deidentified EGRP cohort for manual verification of information such as the age of seizure onset and age of genetic diagnosis. The formatting of these records, however, made running them through our natural language processing (NLP) pipeline unfeasible.

### Clinical term extraction

All clinical annotations analyzed in this study were derived from the full text of the extracted EMR from both cohorts. The full text of clinical notes was run through a customized form of the Clinical Text Analysis and Knowledge Extraction System (cTAKES) NLP pipeline,^20^ which extracts clinical features in the form of Unified Medical Language System (UMLS) codes and, using local contextual information, tags the codes with modifiers such as negation, conditionality, and subject referral. The NLP algorithm NegEx was integrated into the pipeline to aid in negation detection.^21^ Only positive (the clinical feature was documented as present – we did not take into account features documented as absent), current, non-conditional, and subject-related clinical features were analyzed (**Supplementary Table 1**). All UMLS codes were subsequently mapped to Human Phenotype Ontology (HPO; version 1.2; format-version: 1.2; data-version: releases/2020-10-12) terms using the UMLS dictionary (version: 2021AA).^22,23^ All HPO annotations maintained the original time-stamp of the age of the individual at the time of the original note from which the clinical term was extracted.

In the EGRP cohort, we obtained seizure onset and age of genetic diagnosis through manual chart review, verified by a pediatric epileptologist or genetic counselor specialized in pediatric epilepsy and genetics.

### Mapping clinical information onto developmental intervals

Following clinical feature extraction, processed clinical terms were placed into 3-month age bins according to the age of the individual from 0 to 25.25 years old, resulting in 101 time bins (**Fig. 1**). For example, if an individual had the assigned term “febrile seizure” from a medical record at 7 months, 8 months, and 11 months of age, they would have two assigned febrile seizure terms: one in the 6-9-month age bin and another in the 9-12-month age bin. Our previous work has demonstrated the efficacy of this binning strategy to capture clinical landscapes and trajectories in childhood epilepsies.^18,19^ To capture potentially missed edge cases and partially account for surveillance bias, an additional analysis was performed using cumulative binning, e.g., for the age of 9 months all terms per individual between birth and 9 months would be aggregated. Following binning, we utilized the ontological structure of the HPO to perform propagation, a form of automatic reasoning, to infer more general, related clinical concepts from those explicitly extracted from the EMR, as has been reported previously.^17,18,24^ Each individual’s annotations were subsequently de-duplicated within each age bin.

**Figure 1.**
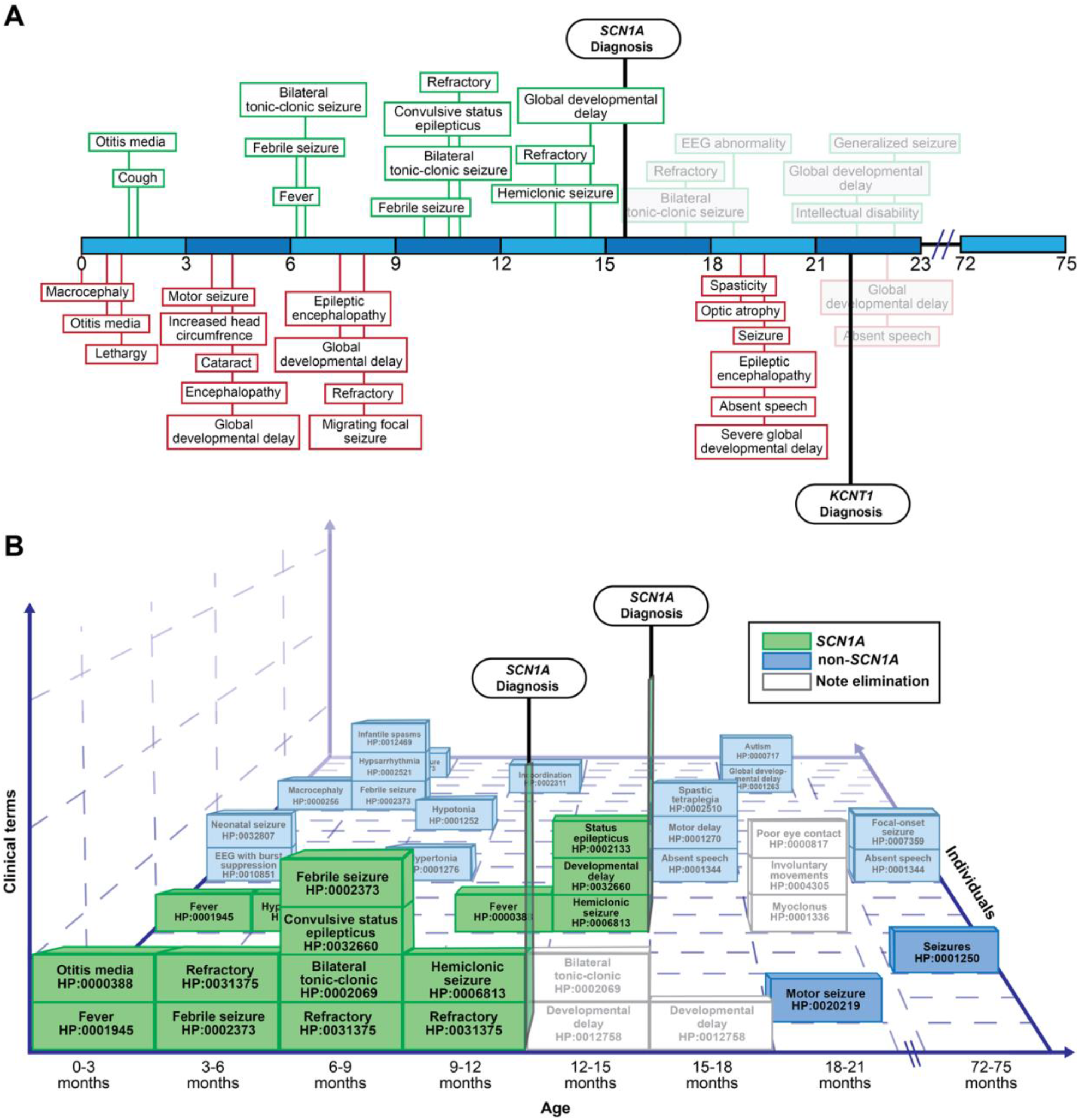
Binning and note elimination. (**A**) A simplified 2-dimensionial representation of how clinical features extracted from the EMR of two patients were placed into 3-month age bins from 0 to 25.25 years for 101 total bins. The top individual, green, was diagnosed with *SCN1A* and the bottom, red, was diagnosed with *KCNT1*. All features within or following an age bin in which an individual received a genetic diagnosis were removed (represented here with faded clinical terms). (**B**) A simplified 3-dimension representation of how these binned terms are grouped according to genetic diagnosis (here, *SCN1A*). Age bins are represented on the x-axis, stacked clinical terms on the y-axis, and individuals on the z-axis. Boxes represent a single clinical term for a particular individual at a particular age bin. Empty, dotted quadrangles signify times at which no clinical features were found for an individual at that time bin.

For many individuals, large gaps in the EMR over time were observed, with correspondingly sparse clinical annotations. These gaps in “EMR usage” may be due to a temporary or permanent resolution of symptoms (e.g., seizure control) but are typically a result of care fragmented across various health systems. For example, many individuals in our cohort with disease-causing variants in *STXBP1* were referred by outside providers to a dedicated specialty clinic, but mainly received care outside our healthcare system. As a result, only small snapshots of their clinical presentation could be retrieved from the EMR. To mitigate potential gaps due to care fragmentation, the general HPO term “All” (HP:0000001) was assigned to every individual in each age bin from their earliest to latest available EMR or until they received a genetic diagnosis (see below).

### Note elimination

We developed a framework to use only clinical notes prior to an individual’s genetic diagnosis. For example, if an individual received a genetic diagnosis of *SCN1A* at 1.3 years of age, all clinical features were removed from the age bin 1.25 to 1.5 years onwards (**Fig. 1**). Edge cases were approached conservatively. Taking the previous example, if the *SCN1A* diagnosis occurred at 1.5 years, the same filtering would occur, removing all clinical features beginning from the age bin 1.25 to 1.5 years. A small subset of the cohort without a known age of genetic diagnosis were removed from primary analyses. The reasoning for this note elimination was two-fold. Firstly, it was the aim of the study to assess clinical features preceding diagnosis in genetic epilepsies. Secondly, note content after the genetic diagnosis is often biased and emphasizes clinical features consistent with the diagnosis, thus making it difficult to draw a clear distinction between clinical impression and genetic diagnosis.

### Isolating significant clinical associations

Individuals were grouped by genetic diagnosis and broader categories of genetic etiologies. For example, an individual with a confirmed causative *de novo* mutation in *SCN1A* would be grouped with other individuals with disease-causing variants in *SCN1A* and with individuals with variants in gene-related groups including, voltage-gated ion channels, sodium channels, and, more broadly, ion channels. Gene group categories were determined using Gene Ontology (see **Supplementary Table 2** for group breakdown).^25,26^ We performed an additional analysis examining all individuals with any genetic diagnosis. This included genetic diagnoses present in only one individual, previously excluded from clinical association analyses.

Unless otherwise stated, analyses were carried out on the combined EGRP and PELHS cohort. Analyses performed exclusively on EGRP were performed primarily for comparison to the expanded cohort (**Table 2**; **Fig. 3B**) and to examine the clinical utility of identified features in the larger undiagnosed PELHS cohort (**Fig. 5**).

### Clinical resemblance to a genetic diagnosis in undiagnosed individuals

To assess the feasibility of employing our results in a clinical setting, we applied our findings of significant clinical features from the EGRP cohort to the largely undiagnosed PELHS population. Significant clinical concepts and their accompanying time-stamped genetic associations were mapped to the reconstructed clinical histories extracted from the EMR of individuals in PELHS.

Here, we applied a more conservative filter, examining only neurological features with significant association with a gene (*P*<0.05), an odds ratio (OR) with 95% confidence interval (CI) with a lower bound >1, and presence in more than one individual with the respective gene and feature in a given time bin. We also took into account the potential effect of the unequal density of clinical terms within the structure of the HPO. For example, “Autism” (HP:0000717) has the broader parent term “Autistic behavior” (HP:0000729). Thus, if a child with *SCN2A* was assigned the term “Autism”, through propagation they would also be assigned the term “Autistic behavior”. If in a particular age bin, both terms exist exclusively in individuals with an eventual *SCN2A* diagnosis, both terms would be statistically identical and redundant. While knowledge of both terms’ significance can be useful clinically, in this analysis it could unfairly strengthen an individual’s association with said genetic diagnosis. Consequently, we applied a method we refer to as “pruning” in which immediate, statistically identical parent terms are removed.^27^

The resulting filtered terms were then grouped by their associated gene and the OR summed. As a result, for every individual at every time bin, the extracted clinical features could be converted to quantities representing the proportional odds of a particular genetic diagnosis. From this approach, we can attempt to effectively map individuals’ phenomes to predict genomes directly from their EMR.

### Statistical analysis

The significance of association between each gene or group of genes and a clinical feature was determined using a two-sided Fisher’s exact test at every three-month age bin. The predictive strength of the clinical feature for the gene or gene class findings remains on a descriptive level and was assessed via the OR with 95% CI and positive predictive values (PPV). The Haldane-Anscombe correction was applied to adjust for an OR of infinity.^28^ Statistical significance was determined at *P*<0.05. Distributions were tested for normality via visual inspection and a Shapiro-Wilk test. Where applicable, *n* refers to the number of individuals with the described genetic diagnosis or class of diagnoses and clinical term in that age bin, i.e., the upper-left quadrant of the contingency matrix. All statistical analyses were performed using the R Statistical Framework.^29^

### Data availability

Primary results are available at cube3.helbiglab.io. Data in a de-identified format will be made available by request to the corresponding author. Code for the primary analyses is available at: https://github.com/galerp/Cube3.

## Results

### Causative genetic variant seen in 38% of individuals with known or presumed genetic epilepsy

Within EGRP (*n*=1,925), we identified causative genetic variants in 738 individuals with 271 unique genetic diagnoses and 87 occurring in two or more individuals. The most common genetic diagnoses were *STXBP1* (*n*=78), *SCN1A* (*n*=55), *SCN2A* (*n*=32), *KCNQ2* (*n*=22), and *CACNA1A* (*n*=18; **Fig. 2**; **Supplementary Table 3**). Through manual chart review, we retrieved age of seizure onset for 1,659/1,822 individuals with seizures (median 2.0 years [IQR 0.58-5.31 years]) and age of genetic diagnosis for 710/738 individuals (median 2.9 years [IQR 1.10-8.48 years]; **Table 1**)

**Figure 2.**
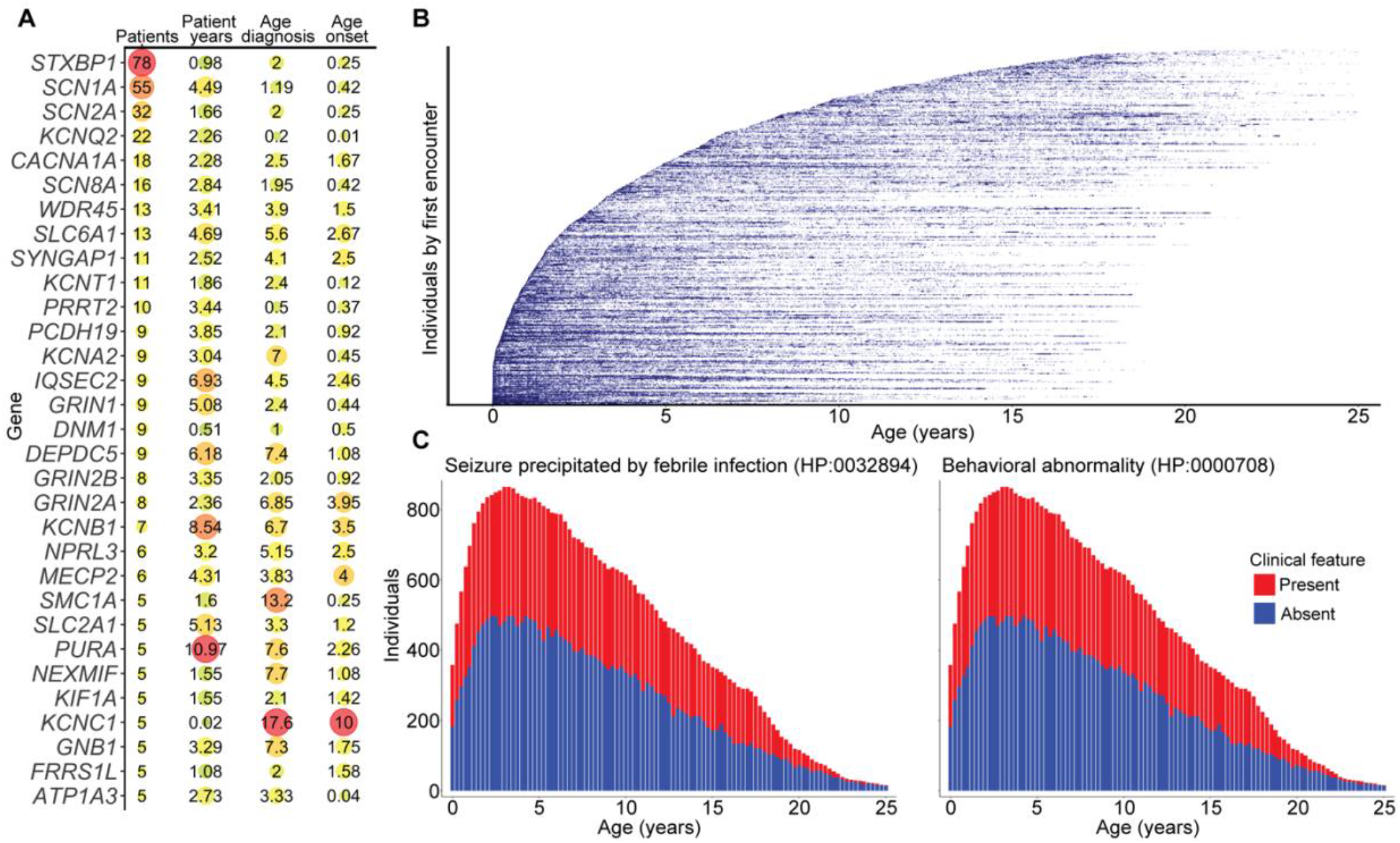
Primary results and cohort representation in the EMR. (**A**) Number of individuals, median patient years in the EMR, median age of genetic diagnosis, and median age of seizure onset of individuals with the respective genetic diagnosis. Colors represent relative size within the respective measurement. For conciseness, only genes with five or more individuals are shown here. (**B**) Every clinical note within the EMR (*n*=689,770) of the EGRP cohort represented as a single data point. X-axis indicates age at a note, while individuals are stacked on the y-axis and sorted by age of earliest note. (**C**) The frequency of clinical terms “seizure precipitated by febrile infection” (HP:0032894) and “behavioral abnormality” (HP:0000708) across development in 3-month time bins displayed as stacked bar charts.

**Table 1.** Demographic information from the EGRP (*n*=1,925) and PELHS (*n*=30,187) cohort.

|  | **EGRP (*n*=1,925)** | **PELHS (*n*=30,187)** |
| --- | --- | --- |
| **Sex** |  |  |
| Male | 1,015 (52.7%) | 16,771 (55.6%) |
| Female | 910 (47.3%) | 13,415 (44.4%) |
| **Race** |  |  |
| Black or African American | 244 (12.7%) | 7,629 (25.3%) |
| Asian | 73 (3.8%) | 1,275 (4.2%) |
| White | 1,201 (62.4%) | 16,391 (54.3%) |
| **Ethnicity** |  |  |
| Hispanic or Latino | 250 (13.0%) | 2,915 (9.7%) |
| Not Hispanic or Latino | 1,620 (84.2%) | 25,171 (83.4%) |
| **Median encounter age (years)** | 6.1 (IQR 2.6–11.6) | 6.6 (IQR 2.7–12.2) |
| **Median age at last follow-up (years)** | 9.0 (IQR 4.9–14.6) | 10.9 (IQR 5.9–16.4) |
| **Median follow-up duration (years)** | 4.5 (IQR 1.8–19.0) | 5.7 (IQR 2.0–10.1) |
| **Median age onset (years)** | 2.0 (*n*=1,659, IQR 0.6–5.3) | -- |
| **Intellectual disability or developmental delay<sup>a</sup>** | 1,130 (58.7%) | 13,405 (44.6%) |
| **Genetic diagnosis** | 738 (38.3%) | -- |
| **Median age diagnosis (years)** | 2.9 (*n*=710, IQR 1.1–8.5) | -- |

<sup>a</sup>PELHS intellectual disability or developmental delay was determined if the individual had the clinical concept neurodevelopmental delay (HP:0012758) and/or intellectual disability (HP:0001249) at any point in their EMR after 1 year of age.

### Childhood epilepsy displays robust presence in EMR

We retrieved all available full-text clinical notes in our healthcare system of every individual in both cohorts. This resulted in 689,770 notes and 10,911 total patient years (median 4.45 [IQR 1.79-8.82]) in the EGRP cohort (**Fig. 2B**) and 3,883,013 notes and 192,458 total patient-years (median 5.67 [IQR 2.02-10.13]) in the PELHS cohort (**Fig. 3A**). EMR usage, the timespan covered by the earliest and latest encounters in the EMR, varied by age. After filters, the age bin in which the greatest number of individuals had annotations was 3.25 to 3.5 years-of-age in EGRP (*n*=866) and 3 to 3.25 years in PELHS (*n*=13,125).

**Figure 3.**
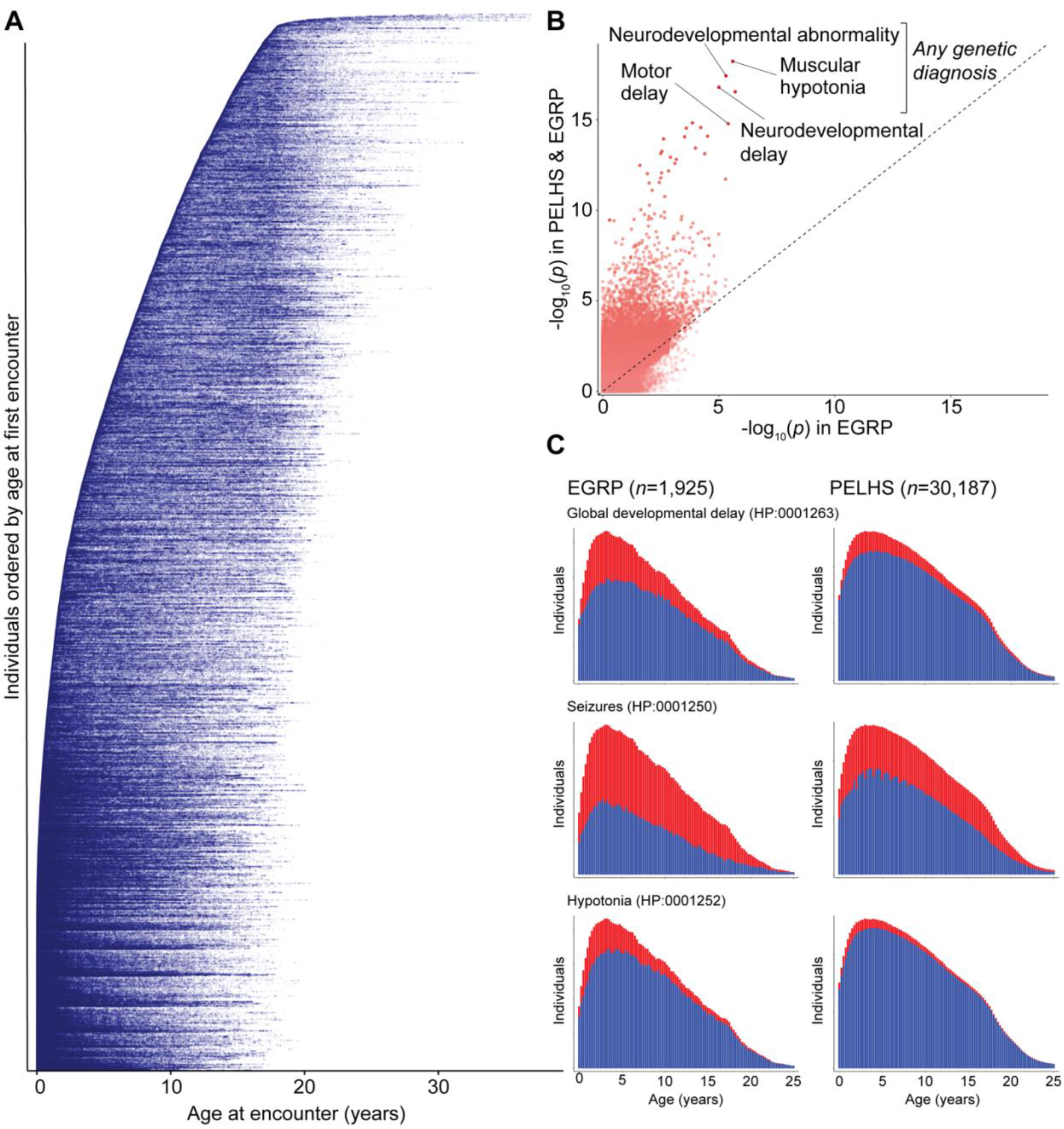
PELHS cohort’s robust representation in the EMR and resulting increase in power. (**A**) Every individual encounter within the EMR of the PELHS cohort below 37 years of age (*n*=1,771,972) represented as a single blue dot. X-axis indicates age at an encounter, while individuals are stacked on the y-axis and sorted by age of earliest encounter. (**B**) Difference in the -log_10_ of the *P*-value of clinical features associated with a gene or gene group prior to genetic diagnosis in the EGRP cohort alone (x-axis; secondary analysis) and the combined EGRP and PELHS cohort (y-axis; primary analysis). The largest gain in significance was found in features associated with any individual with a genetic diagnosis. (**C**) Comparing the prevalence of clinical features across development between the EGRP and PELHS cohort. X-axis is age of individuals in 3-month bins and y-axis is number of individuals.

We retrieved 4,277 and 6,287 distinct clinical features and over 9.3 million and over 80 million clinical annotations in EGRP and PELHS, respectively. False positives were compounded in notes of individuals with a genetic diagnosis, particularly if the diagnosis was disclosed to the individual at the visit of the respective note. We corrected for these errors through filtration via cTAKES modifier tags, reviewing a subset of patient notes with seemingly unrelated clinical findings, and finally through note elimination after genetic diagnosis. After filtering for modifier tags, this resulted in 4,217 and 6,175 distinct HPO concepts annotated 7.3 million and over 61 million times in EGRP and PELHS, respectively.

Despite both cohorts consisting of children with epilepsy from the same hospital system, as seen in **Fig. 3C**, they displayed marked differences in overall clinical presentations across the lifespan. More severe clinical concepts, such as global developmental delay and hypotonia, have a much higher prevalence in the EMR of EGRP individuals throughout their lifespan. Even seizures, a prerequisite for inclusion into PELHS, were annotated over 1.5-fold more in most age bins in EGRP (median=1.57-fold, [IQR 1.33-1.71]). This striking difference indicates the sharp contrast between the often more severe presentation of children with a presumed genetic epilepsy and that of any child presenting with a seizure during their lifespan. These data represent the largest accumulation of clinical EMR-based information from a pediatric epilepsy cohort.

### Earliest features identified a median of 3.6 years prior to median age of diagnosis

Reconstruction of phenotypic profiles in individuals with epilepsy allows for the identification of potentially significant clinical findings detectable prior to the determination of an individual’s genetic cause. We identified a total of 14,440 neurological and 33,201 non-neurological significant clinical features (*P*<0.05, lower 95% OR CI>1; **Table 2**) at distinct ages in 78 of 87 genetic diagnoses present in two or more individuals with extracted clinical concepts after filtering for tagged modifiers (e.g., negation). We found no clinical associations for 9 genetic disorders, as all clinical concepts were completely removed after elimination of notes prior to genetic diagnosis. Of note, 8 of the 9 diagnoses would have clinical terms to analyze if edge cases were permitted (e.g., if an individual with a genetic diagnosis at 12 months of age would retain their clinical terms within the 9-to-12-month age bin).

**Table 2.**
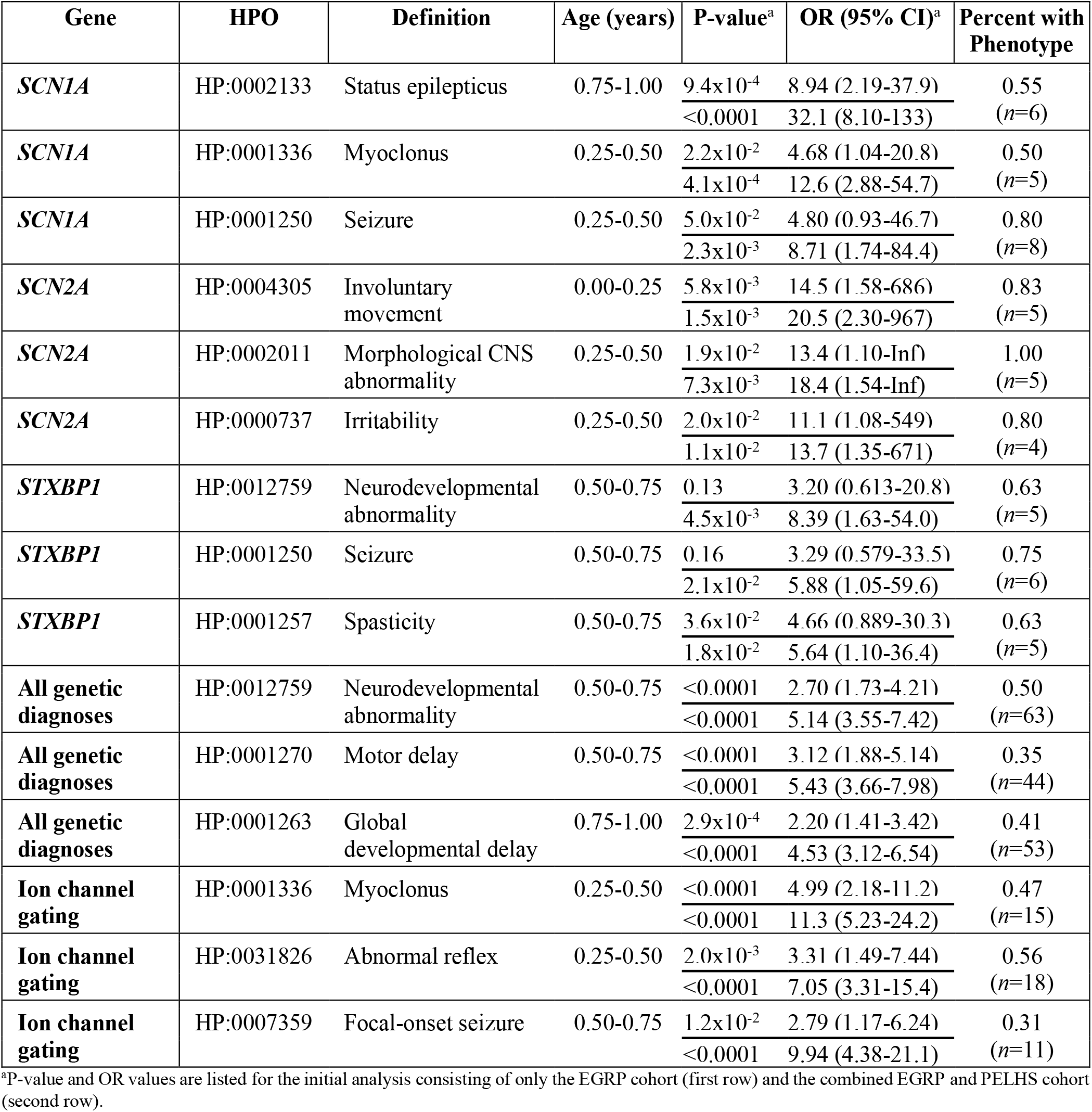
Top clinical features significantly associated with genetic causes and classes prior to diagnosis.

Of the 78 genes with significant clinical findings, the earliest identifiable clinical feature associated with a genetic diagnosis occurred a median of 3.6 years prior to the median age of genetic diagnosis. The earliest neurological features, defined as all clinical concepts subsumed by the “Abnormality of the nervous system” (HP:0000707) branch of the HPO, occurred a median of 2.6 years prior to median age of genetic diagnosis. The longest gaps between earliest significant feature and median age of genetic diagnosis were 13.5 years prior to diagnosis for *MACF1* with cerebellar dysplasia between 7.75 to 8 years (*P*=2.4×10^−4^, 95% CI=107-Inf, PPV=0.33), 13.0 years for *SMC1A* with immunodeficiency at 0 to 3 months (*P*=1.5×10^−2^, 95% CI=1.63-Inf, PPV=0.008), and 10.4 years for *ARFGEF1* with hypertrichosis at 4 to 4.25 years of age (*P*=8.1×10^−3^, 95% CI=3.12-1.60×10^4^, PPV=0.018).

Several genetic diagnoses did not show significant association prior to the median age of genetic diagnosis, including: *TBR1* (*n*=2), 22q11.2 deletion (*n*=2), *TBCK* (*n*=4), and *KCNQ2* (*n*=22). Notably, *KCNQ2* diagnoses typically occurred in early infancy (median=3 months), thus most individuals’ clinical features were completely removed from the primary analyses (*n*=14, 64%).

### Clear clinical associations at least 12 months prior to genetic diagnosis identified in 60 genes

There were 60 genes with more than 10 distinct associated clinical features at least 12 months prior to the median age of genetic diagnosis, including *DDX3X* (1,470 features) with macrotia at 0 to 3 months of age and 4.85 years prior to median diagnosis (*P*=6.4×10^−4^, 95% CI=40.3-Inf), *SLC6A1* (378 features) with retrocollis at 0 to 3 months of age and 5.35 years prior to median diagnosis (*P*=1.0×10^−3^, 95% CI=25.1-4.50×10^15^), and *SCN3A* (1,394 features) with polymicrogyria at 9 to 12 months of age and 10.20 years prior to median diagnosis (*P*=7.0×10^−3^, 95% CI=3.59-Inf).

Other genes present in ten or more individuals (**Fig. 2A**) had clear, significant features prior to genetic diagnosis. Notable among these were *CACNA1A* with agenesis of corpus callosum at 3 to 6 months (*P*=1.7×10^−3^, 95% CI=4.31-Inf) preceding median genetic diagnosis by 2.00 years, *SYNGAP1* with muscular hypotonia at 1 to 1.25 years (*P*=9.9×10^−4^, 95% CI=3.74-Inf) preceding median diagnosis by 2.85 years, and *KCNT1* with epileptic encephalopathy at 3 to 6 months (*P*<0.0001, 95% CI=35.0-Inf) preceding median diagnosis by 1.90 years. Several strong associations were found with genes with five individuals or fewer: *CDKL5* with myoclonus between 0 to 3 months (*P*=8.9×10^−3^, 95% CI=1.80-Inf) preceding median diagnosis by 1.45 years, *SMC1A* with microcephaly between 3 to 6 months (*P*=7.9×10^−4^, 95% CI=6.51-Inf) preceding median diagnosis by 12.70 years, and *PURA* with muscle weakness between 9 to 12 months (*P*=7.8×10^−4^, 95% CI=6.54-Inf) preceding median diagnosis by 6.60 years.

### Well-established clinical associations found in undiagnosed individuals

Notably, many of our findings reflect well-established clinical associations, nevertheless were detected in as of yet undiagnosed individuals. For example, between the age of 6 and 9 months, four individuals with a diagnostic *SCN1A* variant presented with status epilepticus with prominent motor symptoms (*P*<0.0001, 95% CI=19.5-510; **Fig. 4**). Of these infants, three had not been diagnosed with *SCN1A* by 9 months of age and would not be diagnosed for nearly one year. Similarly, four individuals with *de novo* mutations in *STXBP1* had encephalopathy between 0 and 3 months (*P*=7.1×10^−4^, 95% CI=3.99-2.57×10^3^). Three of these neonates had yet to be diagnosed with *STXBP1*, two of which would not receive this diagnosis for at least 9 months.

**Figure 4.**
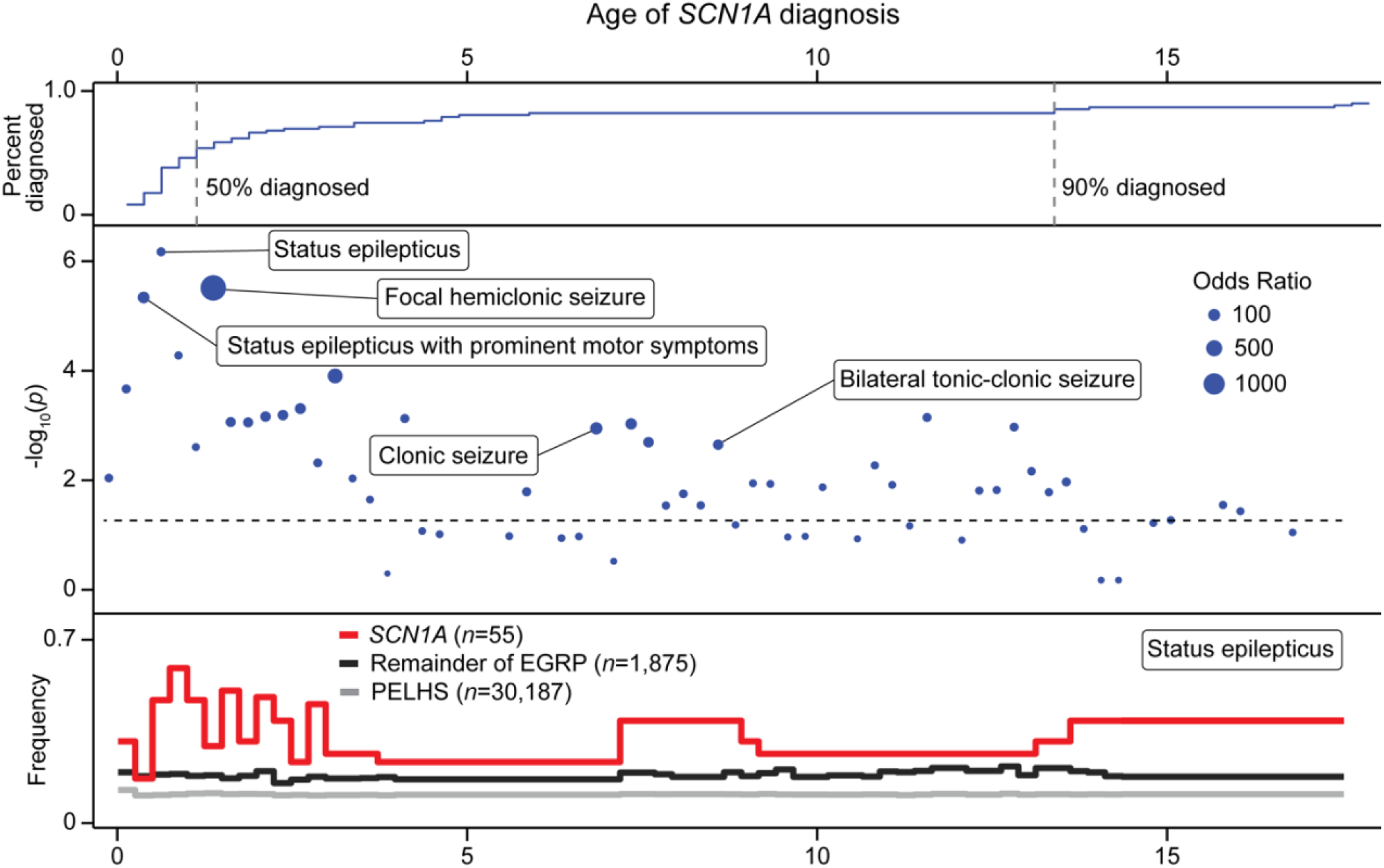
Significant clinical features in *SCN1A* across age bins prior to genetic diagnosis. The top panel depicts the cumulative proportion of individuals receiving a genetic diagnosis of *SCN1A*. The middle panel displays the most significant clinical features prior to genetic diagnosis in each 3-month time bin. Labeling was determined based on clinical interest and legibility (other terms can be seen via our web interface). X-axis denotes patient age, y-axis denotes -log_10_ of the *P*-value, size of point indicates relative odds ratio (OR), and the dotted line indicates -log_10_(0.05). The bottom panel represents the frequency of status epilepticus prior to genetic diagnosis in *SCN1A*, individuals in the EGRP cohort without a *SCN1A* diagnosis, and the PELHS cohort.

### Novel clinical features found in several genetic epilepsies

While confirming known early clinical features in genetic epilepsies, our study also identified novel associations. This included associations such as *IQSEC2* with aplasia/hypoplasia involving the central nervous system, most significant between 3 and 3.25 years (*P*=3.3×10^−4^, 95% CI=5.24-3.35×10^3^, *n*=3), *SCN2A* with yellow/white lesions of the macula, most significant between 1.5 to 1.75 years (*P*=3.1×10^−3^, 95% CI=2.33-1.51×10^3^, *n*=3), *DEPDC5* with abnormal fear/anxiety-related behavior, most significant between 5.75 and 6 years (*P*=5.5×10^−4^, 95% CI=4.36-2.80×10^3^, *n*=3), and *KCNT1* with abnormality of the kidney, most significant between 9 and 12 months (*P*=8.9×10^−4^, 95% CI=3.88-Inf, *n*=3).

### Gene groups display significant clinical overlap

Many of the identified genes are essential components of similar neurophysiological processes and pathways. Grouping these genetic diagnoses into these broader classes (**Supplementary Table 3**) revealed additional associated clinical features prior to individuals’ diagnoses. Variants in genes involved in ion channel gating showed significant associations with myoclonus (*P*<0.0001, 95% CI=5.23-24.2, *n*=15) and seizures between 3 to 6 months (*P*<0.0001, 95% CI=2.49-13.8, *n*=23). Variants in sodium-channel-related genes were associated with clonic seizures between 3 to 6 months (*P*<0.0001, 95% CI=5.09-86.2, *n*=5) and status epilepticus between 1 to 1.25 years (*P*<0.0001, 95% CI=2.15-112, *n*=7). Between 6 to 9 months of age, variants in ligand-gated channel-related genes were associated with global developmental delay (*P*<0.0001, 95% CI=10.7-Inf, *n*=7) and glutamate-receptor-related genes were associated with motor delay (*P*<0.0001, 95% CI=9.00-Inf, *n*=5).

### Broad clinical features are highly predictive of achieving a genetic diagnosis

We hypothesized that certain clinical features may be predictive of eventually receiving a genetic diagnosis in children with epilepsy. Consequently, we examined clinical terms associated with any genetic diagnosis, assessing clinical features linked to genetic epilepsies more generally. Terms with the strongest associations included muscular hypotonia between 1 to 1.25 years (*P*<0.0001, 95% CI=4.14-8.73, *n*=51), neurodevelopmental abnormality between 6 to 9 months (*P*<0.0001, 95% CI=3.55-7.42, *n*=63), and neurodevelopmental delay between 6 to 9 months (*P*<0.0001, 95% CI=3.51-7.33, *n*=60). These findings represent early clinical features indicative of receiving diagnostic results following genetic testing. Of note, the strength of clinical terms associated with the broader group of any genetic diagnosis became substantially more prominent than associations with any other genes or gene groups following the addition of the PELHS cohort to the analyses (**Fig. 3B**). The clinical associations with the genetic diagnoses and other relevant results within this study can be searched and visualized online (**cube3.helbiglab.io**).

### Cumulative age bins reveal hidden clinical features

Discrete age binning can potentially obscure relevant clinical information, particularly in genetic disorders with few individuals and sparse data. The known association of migrating focal seizures with *KCNT1* became apparent in our data only when examining bins cumulatively: at 0 to 9 months (*P*<0.0001, 95% CI=174-Inf, PPV=0.17) and continuing to 0 to 1.75 years (*P*<0.0001, 95% CI=96.8-4.50×10^15^, PPV=0.13). The analysis also revealed additional significant associations, such as between *ARFGEF1* and generalized non-convulsive status epilepticus without coma from 0 to 10 years (*P*<0.0001, 95% CI=51.6-Inf, PPV=0.021) up to 12 years (*P*<0.0001, 95% CI=43.3-Inf) and *GRIN2A* and slurred speech from 0 to 5 years (*P*<0.0001, 95% CI=47.3-Inf, PPV=1.6×10^−5^) up to 5.25 years (*P*<0.0001, 95% CI=43.2-Inf, PPV=1.9×10^−5^). Furthermore, the windows of associations from previous analyses were extended such as *SCN1A* with status epilepticus from 0 to 9 months (*P*=1.4×10^−3^, 95% CI=2.11-31.1, PPV=6.0×10^−3^) up to 4.25 years (*P*=1.1×10^−2^, 95% CI=1.32-102, PPV=1.2×10^−3^) and *STXBP1* with epileptic encephalopathy from 0 to 1 year (*P*=4.3×10^−3^, 95% CI=2.55-199, PPV=4.3×10^−3^).

### Determining likely genetic diagnoses in the undiagnosed

Next, we examined the clinical resemblance to genetic epilepsies across the age span of undiagnosed individuals (**Fig. 5**). We found that the most likely genetic diagnosis for an individual often changes across the lifespan. For example, subject RE56AO3I600FY entered the CHOP care network at 3.48 months of age, presenting with features strongly predictive of *SCN1A* including febrile seizures (*P*=2.4×10^−2^, 95% CI=1.02-30.3, PPV=0.10) and clonic seizures (*P*=1.8×10^−2^, 95% CI=1.15-34.8, PPV=0.12). As seen in **Fig. 4**, with advancing age, associations with clinical features weaken in genes such as *SCN1A* and *SCN2A* that typically cause early-onset presentations, as opposed to genes such as *DEPDC5* with later presentations. We found that other individuals, such as RE5AF5VV7SJIDM, RE5LLT7594M9F, and RE55KEWJAZVZ4, with prominent early clinical features display strong resemblance to a few genetic diagnoses such as *KCNT1, SCN1A*, or *SCN2A*, suggesting that these individuals might benefit from genetic testing.

**Figure 5.**
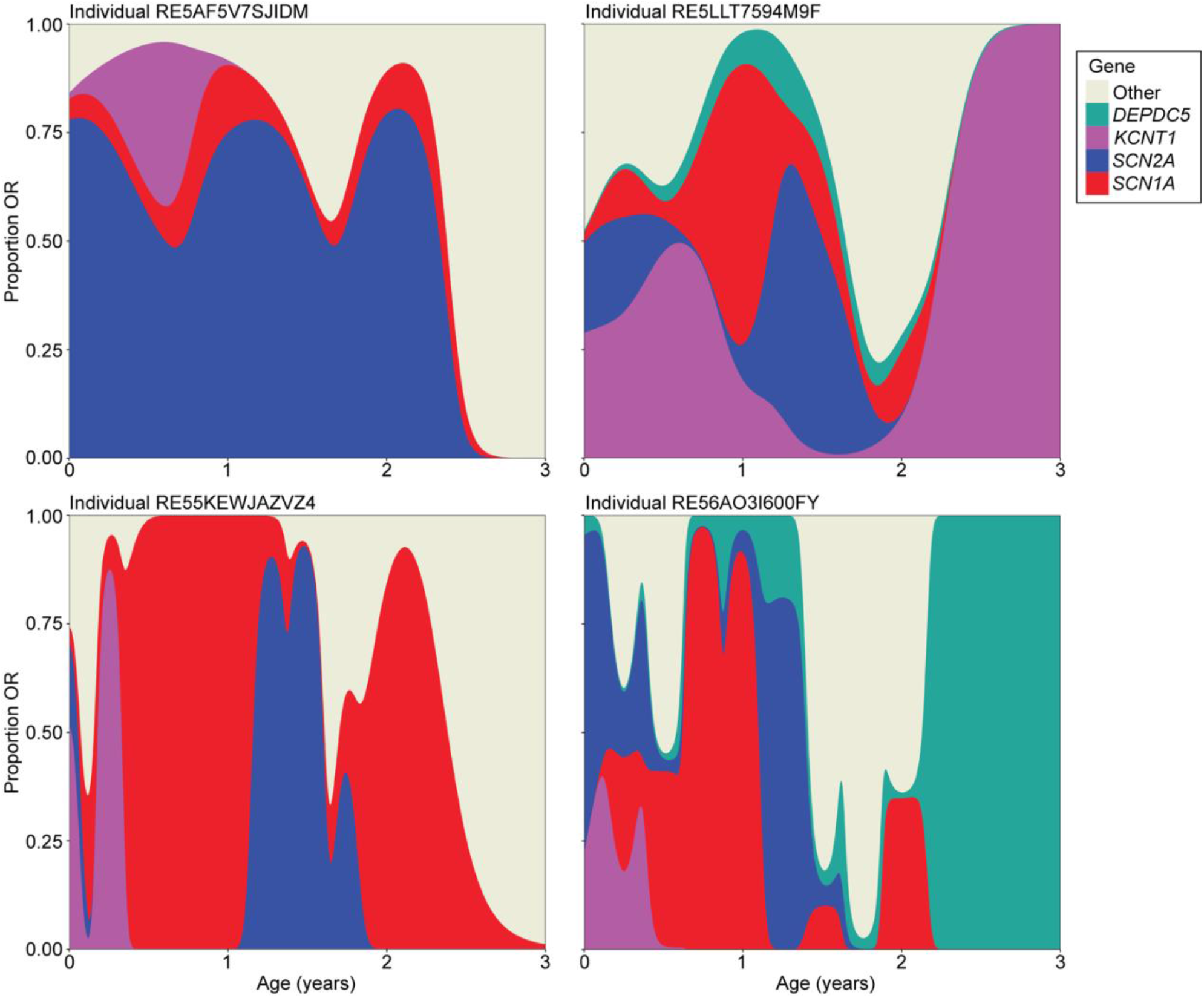
Clinical similarity to a genetic cause in undiagnosed patients across their development. Four individuals within the PELHS cohort with their predicted genetic diagnosis across development according to extracted clinical features at a particular age. The x-axis is age in years and the y-axis is the proportion of the total summed odds ratio (OR) of significant clinical terms associated with a particular genetic diagnosis. OR proportion was smoothed across 1 year to demonstrate broader patterns. “Other” signifies every genetic diagnosis besides those listed.

### Characteristics of individuals with prolonged diagnostic delay

We next sought to determine characteristics in individuals that may indicate a more prolonged delay in receiving a genetic diagnosis. To estimate this “diagnostic delay,” we subtracted the age of seizure onset from the age of genetic diagnosis for those with both data points available in EGRP (*n*=575). While imperfect, seizure onset provides an approximation for most individuals of when notable, adverse symptoms of their epilepsy disorder began. Individuals were split by the severity of their delay in receiving a diagnosis: between those below (<1.20 years; moderate delay; *n*=285) and those above and equal to the second quartile (**>=**1.20 years; severe delay; *n*=290).

We hypothesized that the severity of diagnostic delay was due in part to insufficient access to genetic testing. First, we hypothesized that age of the patient would be a factor, as older individuals would be more likely to undergo testing further in the past and therefore not have benefited from more affordable, advanced, or modern genetic testing and variant interpretation. Upon visual inspection and a Shapiro-Wilk test, the age distributions were non-parametric. Thus, we applied a two-sample, one-sided Kolmogorov-Smirnov test, finding a significant difference in distributions between the individuals with moderate and severe diagnostic delay (*P*<0.0001), indicating those with greater delay were typically older. The type and trajectory of genetic tests performed displayed similar results.

Those with severe diagnostic delay were more likely to have undergone single-gene testing (*P*=5.5×10^−3^, 95% CI=1.23-3.35, *n*=57), but eventually receive a final genetic diagnosis from whole-exome or whole-genome testing (*P*<0.0001, 95% CI=1.88-3.93, *n*=150). This group was also significantly enriched for individuals with rarer genetic diagnoses, here defined as genes that appeared in 2 or less individuals in the EGRP cohort (*P*=2.7×10^−4^, 95% CI=1.34-2.81, *n*=116). This was marginally significant when considering genetic diagnoses that appeared only once in the cohort (*P*=3.6×10^−2^, 95% CI=1.03-2.36, *n*=76). Together this suggests that if the individuals had earlier access to more comprehensive genetic testing, they would have received a genetic diagnosis sooner. As the age-related differences in diagnostic delay demonstrates, however, we expect that this delay may be naturally reducing due to expanding access to more advanced and inexpensive genetic testing.

## Discussion

In our study, we identify early clinical features associated with a genetic cause prior to diagnosis, using the rich information available in the EMR. Genetic epilepsies represent a growing proportion of childhood epilepsies with increasing focus on precision therapies and early diagnosis. An increasing number of genetic epilepsies have genetically targeted treatment and management.^30,31^ However, early diagnosis traditionally requires expert knowledge, which becomes a bottleneck when large patient populations need to be served. Accordingly, there is a critical need for tools to prioritize patients for genetic testing that do not rely on expert determination. One method to accomplish this is by employing frameworks that mine existing medical records, such as the clinical information already in EMR.^18,19^

### A novel method for gene characterization

Our approach to using EMR data for gene characterization differs significantly from prior tools. Most solutions to prioritize genetic diagnosis from clinical information have generated a gene ranking system.^32-34^ Existing tools largely use static, manually submitted data to predict genetic diagnoses based on clinical features listed in reference databases such as OMIM, which are based on published cases and unlikely to reflect real-world prevalence of phenotypes. Moreover, unlike the pipeline presented here, there is no distinction in the age when the clinical feature presents or abates. For example, most existing tools would not differentiate between status epilepticus in an individual at 1 week and 15 years of age.^32-34^

While well-established, as with all NLPs, the cTAKES pipeline is imperfect, particularly in detecting phenotypes that are erroneous or absent in clinical notes. For example, pneumocystis jirovecii pneumonia is a rare condition with no known correlation to epilepsy disorders, yet it was detected in more than 80% of individuals from the EGRP cohort (*n*=1,551). Upon further examination, the cTAKES pipeline was identifying this phenotype from the string “PCP”, a common acronym for “primary care physician” that can also be used to refer to this type of pneumonia. Such errors were corrected to the best of our ability through manual review.

There has been recent work demonstrating that more advanced NLP exploiting deep learning can extract detailed information such as seizure frequency from the EMR of individuals with epilepsy more accurately.^35^ Integrating such tools could provide a more accurate, deeper picture of an individual’s clinical landscape.

### Leveraging the EMR from 32,112 individuals with epilepsy

We reasoned that the standardized clinical concepts extracted from approximately 4.6 million full-text clinical notes from 1,925 individuals with known or presumed genetic epilepsies could reveal early indicative features, particularly when compared to 30,187 other individuals with epilepsy from a large pediatric health network. We found significantly associated features prior to genetic diagnosis in 78 of the 87 genes analyzed, providing new insights into the early clinical trajectories of genetic epilepsies, potentially allowing for automated screening in routine care.

Even when clinical features in genetic epilepsies are known, it is often difficult to assess how these features compare to other genetic diagnoses specifically or to childhood epilepsies in general. For example, infantile spasms are present in 60% of individuals with *STXBP1*-related disorders before 3 months of age, but only in 6.2% of individuals with other known or presumed genetic epilepsies and in 1.5% of a large pediatric epilepsy cohort at that age period. This represents a 40-fold increased risk for an individual with infantile spasms to have a genetic diagnosis of an *STXBP1*-related disorders. Likewise, we demonstrated that status epilepticus between 6 and 9 months is predictive of a molecular diagnosis of an *SCN1A*-related disorder, anticipating the current median diagnostic age by over 3 years.^2^ At this age, this feature was present in 66% of individuals with an *SCN1A*-related disorder, 18% of individuals with other known or presumed genetic epilepsies, and only in 3.2% of the larger pediatric epilepsy cohort present in the EMR at this age span. With a 21-fold increased risk for an *SCN1A* diagnosis, systematically screening individuals presenting with this feature may be warranted. Within our larger epilepsy cohort, this would have applied to a median of 667 (IQR 581-691) children per month across all ages between January 2020 and May 2022 (**Supplementary Fig. 1**).

### Gene groups and all genetic diagnoses display robust clinical associations

Given the rarity of individual genetic diagnoses, we assessed effects across gene classes and all genetic epilepsies combined, finding that associations tended to strengthen when considering gene groups (**Fig. 3B**). For example, ion-channel-related genes were associated with motor seizures between 3 to 6 months (*P*<0.0001, 95% CI=3.43-17.8) and ligand-gated-channel-related genes were associated with muscular hypotonia between 6 to 9 months (*P*<0.0001, 95% CI=6.93-2.59×10^3^). Such results provide an impetus for further work expanding beyond the gene-specific approach to more broad groups of genes with overlapping pathophysiological pathways.

Furthermore, if an individual with epilepsy presents with relatively non-specific clinical features such as muscular hypotonia, neurodevelopmental delay, or neurodevelopmental abnormality between 6 to 15 months, a subsequent genetic diagnosis is 4-6 times more likely. With positive predictive values of many features for a later genetic diagnosis exceeding 10%, the theoretical diagnostic yield of our methodology approaches the yield for gene panel testing, which is estimated to be 10-20% in childhood epilepsies.^31,36,37^

The strength of these features within gene groups indicates that many genetic epilepsies have overlapping, yet specific clinical features beyond recognizable individual conditions such as *SCN1A*-related epilepsies. Moreover, individuals who initially underwent narrower genetic testing workups displayed higher latency to diagnosis. These findings justify the current practice of using comprehensive gene panels or exome/genome sequencing as a first line diagnostic test that allows for the identification of any of the potentially causative genes. Furthermore, knowledge of such phenotypic markers in a clinical setting, either directly from the clinician or integrated within the EMR, could improve genetic testing throughput, diagnostic yield, and treatment in infants and young children with epilepsy. To this end, we have provided the clinical associations identified in this study online (**cube3.helbiglab.io**).

### Unconstrained EMR and confirmation bias

In our study, we also attempted to address inherent treachery in EMR data. Particularly in rare diseases, full-text clinical notes often include discussions of the underlying genetic diagnosis which can bias assessment of clinical features. For example, most individuals with *GRIN2A*-related disorders carry both an ICD diagnosis and documented clinical finding of speech apraxia. However, only two of the eight individuals with this diagnosis presented with this feature in their notes prior to their genetic diagnosis.

Similarly, our preliminary analyses revealed skin vesicles to be significantly associated with individuals with a *de novo* mutation in *STXBP1*. This was due to incorrect extraction of a clinical feature from repeatedly used text explaining the genetic diagnosis: “*STXBP1* encodes the syntaxin-binding protein 1, which plays an important role in the regulation of synaptic *vesicle* docking and fusion…” These long strings of text describing a genetic disorder or other diagnosis are often copied and reused among clinicians in their notes. This is often referred to as “copy-forwarding”.

These phenomena, which we refer to collectively as “note contamination,” make it difficult to separate an individual’s clinical course from the genetic diagnosis. Retrospective studies of disease associations, particularly those employing unfiltered EMR, are particularly susceptible to these inherent human and algorithmic biases. We corrected for most of these errors through note elimination after genetic diagnosis. Systematically removing clinical notes after genetic diagnoses enabled us to identify significant, age-dependent clinical features prior to the age of diagnosis in 78 genetic etiologies, highlighting the potential for using this information to achieve earlier diagnoses.

### Limitations

For the purposes of this study, the PELHS cohort in this study is assumed not to have received any epilepsy genetic diagnoses. This assumption could not be verified as the cohort is deidentified. Under EGRP, there has been a concerted effort to recruit those individuals seen at CHOP with a known or presumed genetic epilepsy. Nevertheless, it is possible that some individuals with genetic epilepsies were absent from this subcohort. In many ways, however, this demonstrates the strength of our associations, as any such individuals may lead to lower measured effect sizes and reduced statistical power. For example, if an individual within PELHS had a causative variant in *SCN2A*, any *SCN2A*-related clinical features in their EMR would contribute to a weaker measured association between the feature and *SCN2A* diagnosis, especially because no note elimination would have been carried out following this genetic diagnosis. While this may result in some false negative or missed associations, we would not expect this to cause false positives, so any detected associations are robust to this possible misclassification.

The strength of our findings is also notable because, as our previous work has shown, genetic epilepsies can be highly heterogeneous.^17^ While their clinical presentations tend to demonstrate significant similarity, this is variable and can rise and fall across development.^18,19^ We expect this baseline variability is largely due to factors we cannot account for, including developmental environments, different individual variants, and polygenic interactions. These effects may be exacerbated within our dataset as it originates from a single hospital system with limited representation from underserved populations and few international individuals, a common problem in clinical and research genetics in general.^38,39^ Our cumulative binning strategy, however, helps to partially overcome the potential downstream effects of these factors on delayed or accelerated presentation and documentation of clinical features. Furthermore, it is our hope that studies such as this can promote access to genetic testing in wider populations.

## Conclusion

In summary, we capture clinical features associated with genetic epilepsies years prior to genetic diagnosis, using relatively easily scalable tools and existing medical records. Furthermore, we map the trajectory of phenome-genome relationship across development, tracking the natural history of rare genetic disorders in unprecedented detail. The approach and features outlined in this work can contribute to earlier diagnoses, more precise prognostication, and genetically informed treatment strategies, thereby providing clinicians valuable clinical decision support for the genetic epilepsies in the precision medicine era.

## Data Availability

All data produced in the present study are available upon reasonable request to the authors.

https://eig.research.chop.edu/cube3/

## Abbreviations

CI: Confidence Interval
EGRP: Epilepsy Genetics Research Project
EMR: Electronic Medical Records
HPO: Human Phenotype Ontology
NLP: Natural Language Processing
OR: Odds Ratio
PELHS: Pediatric Epilepsy Learning Health System
PPV: Positive Predictive Value

## Acknowledgements

We thank the participants and their family members for taking part in the study. We would like to thank Mahgenn Cosico and Priya Vaidiswaran for their support in enrolling research participants and for administrative assistance. We thank the Arcus team at Children’s Hospital of Philadelphia for support with data infrastructure and analysis.

## Supplementary material

Supplementary material is available at *Brain* online.

