## Supplement for "Clinical signatures of genetic epilepsy precede diagnosis in electronic medical records of 32,000 individuals"

**Supplementary material**

**Table of Contents**

**Supplementary Definition of PELHS cohort**………………………….…………….....….Page 2

**Supplementary Description of cTAKES pipeline and filters**…………………….........Page 2-3

**Supplementary Results from monthly counts of clinical features**…………....…….........Page 3

**Supplementary Figure 1. Monthly counts of four clinical concepts from PELHS**….….Page 4

**Supplementary Table 1 Filters applied using cTAKES modifier tags**..............................Page 5

**Supplementary Table 2 Gene group compositions from EGRP**........................................Page 6

**Supplementary Table 3 Overview of data on genes from EGRP**...……….....…….......Page 7-8

**References**……………………...…………...…………...…………...….………….............Page 9

**Supplementary Definition of PELHS cohort**

The Pediatric Epilepsy Learning Health System (PELHS) cohort was collected by searching the Children’s Hospital of Philadelphia (CHOP) electronic medical records (EMR) for every individual with who has received a diagnosis of any ICD-10/9 epilepsy code, including: ICD codes: ICD9:345.x, ICD9:779.0, ICD10:G40.x, ICD10:R56.x, or ICD10:P90.x. The search included every in individual under age 18 seen in the CHOP primary health system since 2010. It has been previously published using structured EMR data from the cohort.^1^ Since the EGRP cohort consists of individuals seen at CHOP with epilepsy, most of these individuals are included within PELHS. Consequently, we took steps to remove these patients from PELHS prior to any analyses. Besides those individuals that also exist within EGRP, PELHS is entirely deidentified thus preventing us access to information such as exact dates of encounters, genetic diagnoses, or seizure age of onsets. We were, however, able to obtain year and month counts of extracted clinical features (as seen in **Supplementary Fig. 1**). For these counts, we included individuals from both PELHS and EGRP (removing duplicated patients) to ensure that we could obtain the most accurate and full picture of the landscape of individuals with epilepsy seen within our hospital system.

Because PELHS is entirely deidentified, besides those that also exist within EGRP, we could not collect potential genetic data nor directly read their EMR to check for a genetic diagnosis. Many clinicians and research staff, however, have made a concerted effort since 2014 to identify and consent individuals and their families that have a known or likely genetic mutation. Nevertheless, it is likely that some individuals within PELHS have received a genetic diagnosis and are not consented within EGRP.

**Supplementary description of the cTAKES pipeline and filters**

Apache cTAKES pipeline has been trained on gold standard annotations from the Penn TreeBank (PTB), GENIA corpora, and Mayo Clinic EHR notes achieving F scores up until entity recognition of >0.9 under most measures. It can detect complex phenotype relationships (“negation”, “history of”, “uncertainty”, and “conditional” statements and respective “subject”), and it is modular, allowing for the user to pinpoint areas the pipeline to improve and add additional features.^2^ The pipeline is run at CHOP with Google Cloud Platform (GCP).

To integrate the Human Phenotype Ontology (HPO; version 1.2; release format-version: 1.2; data-version: releases/2021-04-13),^3^ the Entity Recognition module has been altered, mapping the Unified Medical Language System (UMLS) ID output to the equivalent HPO term. UMLS is a collection of dictionaries, files, and software produced to harmonize clinical and biomedical vocabularies, adding to the inherent flexibility of the pipeline.^4^

Apache cTAKES tags extracted clinical concept with the modifiers: negation, conditionality, generic, history-of, uncertainty, and subject referral. The NLP NegEx was added to the pipeline to aid in negation detection.^5^ Below in **Supplementary Table 2** displays how we filtered with these modifiers.

**Supplementary results from monthly counts of clinical features**

As an additional analysis, we were able to pull a monthly count of all clinical features within the CHOP Care Network from January 2010 to May 2022 within all of PELHS (including those also within EGRP). In **Supplementary Fig. 1**, we present some of those results. The general trend exhibits the growth of the CHOP Care Network and its EMR system, with a notable drop during the initial lockdown phase of the 2020 COVID-19 pandemic.

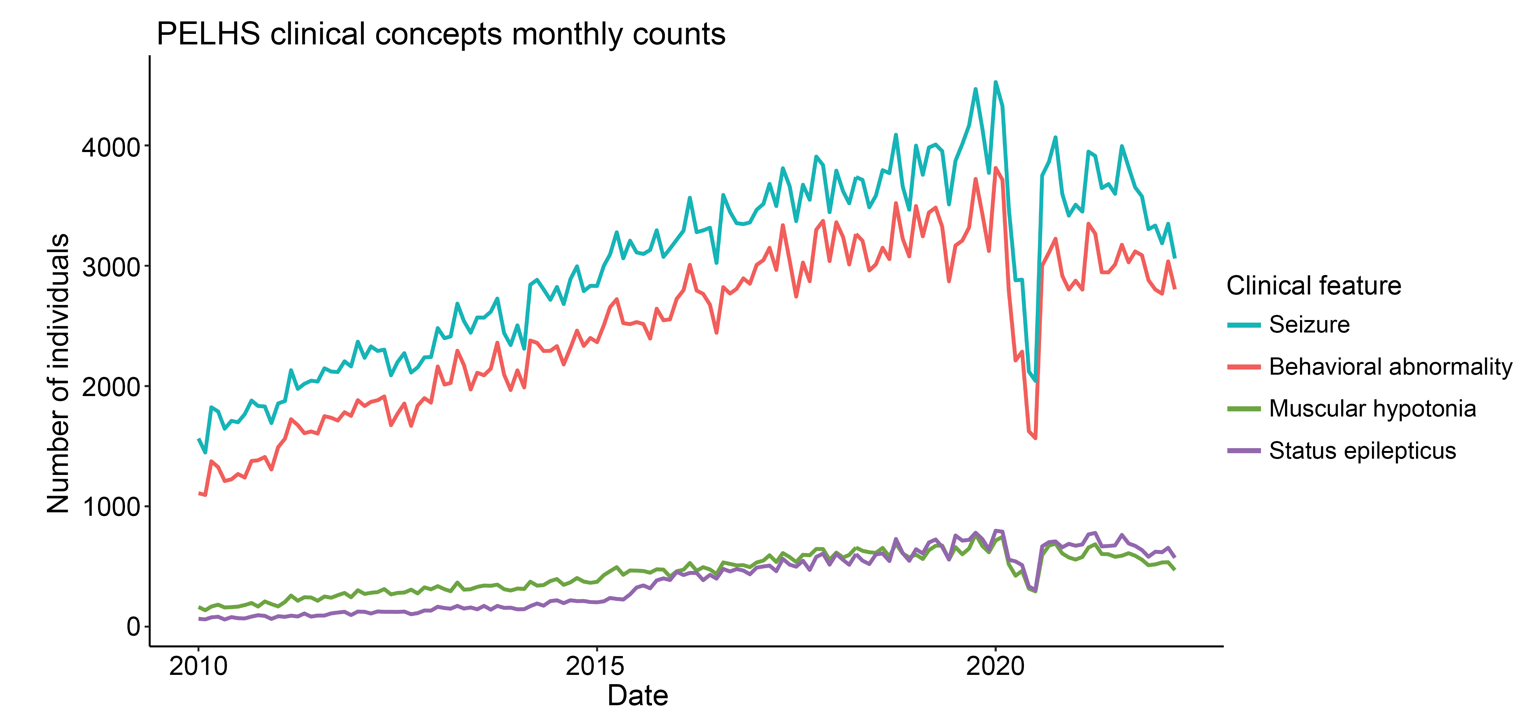

**Supplementary Figure 1 Monthly counts of four clinical concepts from PELHS.** The number of individuals per month from January 2010 to May 2022 within all of PELHS (including individuals also within EGRP) with the respective clinical feature extracted from their EMR.

**Supplementary Table 1 Filters applied using cTAKES modifier tags.**

| **Modifier Tag** | **Possible Tags** | **Filter** | **Example of data excluded** |
| --- | --- | --- | --- |
| Negation | True/False | False | *“[Patient is]* *currently on 1 mL BID for hypsarrhythmia on EEG (****no clinical spasms****)…”* |
| Conditional | True/False | False | *“[Patient] had outpatient…with neuro who stated pt may require depakote* ***if seizures*** *again through keppra…”* |
| Generic | True/False | False | *“She saw [clinician] more recently in clinic and Trileptal was increased at that visit,* ***headaches were mentioned that were nonspecific*** *sounding.”* |
| History-of | True/False | False | *“Pertinent medical* ***history of polymicrogyria, infantile spasms/seizures, adrenal insufficiency, and developmental delays****…”* |
| Uncertainty | True/False | False | *“…the surface of the brain appears smooth with double-cortex appearance that* ***could represent lissencephaly/pachygyria*** *spectrum…”* |
| Subject | Patient/Family-Member/Other | Patient | *“****Family history of mitral valve prolapse****…”* |

**Supplementary Table 2 Gene group compositions from EGRP.**

| **Gene Class** | **Genes (number of individuals)** |
| --- | --- |
| Calcium channels | *CACNA1A* (18); *CACNA1E* (3); *CACNA1C* (1); *CACNA1G* (1) |
| Calcium voltage | *CACNA1A* (18); *CACNA1E* (3); *CACNA1C* (1); *CACNA1G* (1) |
| Gaba receptors | *GABRG2* (4); *GABRB3* (3); *GABRA3* (2); *GABRA1* (1); *GABRB2* (1) |
| Glutamate ionotropic | *GRIN1* (9); *GRIN2A* (8); *GRIN2B* (8); *GRIN2D* (2); *GRIA3* (1) |
| Glutamate receptors | *GRIN1* (9); *GRIN2A* (8); *GRIN2B* (8); *GRIN2D* (2); *GRIA3* (1) |
| Ion channel | *SCN1A* (55); *SCN2A* (32); *KCNQ2* (22); *CACNA1A* (18); *SCN8A* (16); *KCNT1* (11); *KCNA2* (9); *KCNB1* (7); *KCNC1* (5); *CACNA1E* (3); *SCN3A* (3); *KCNA1* (2); *CACNA1C* (1); *CACNA1G* (1); *CLCN4* (1); *KCNMA1* (1); *KCNN2* (1); *KCNK18* (1); *KCNK4* (1); *KCNC2* (1); *KCND3* (1); *KCNQ3* (1) |
| Ion channel gating mechanism | *SCN1A* (55); *SCN2A* (32); *KCNQ2* (22); *CACNA1A* (18); *SCN8A* (16); *GRIN1* (9); *KCNA2* (9); *GRIN2A* (8); *GRIN2B* (8); *KCNB1* (7); *KCNC1* (5); *GABRG2* (4); *CACNA1E* (3); *HCN1* (3); *GABRB3* (3); *SCN3A* (3); *GABRA3* (2); *GRIN2D* (2); *KCNA1* (2); *CACNA1C* (1); *CACNA1G* (1); *GABRA1* (1); *GABRB2* (1); *GRIA3* (1); *KCNMA1* (1); *KCNN2* (1); *KCNK18* (1); *KCNK4* (1); *KCNC2* (1); *KCND3* (1); *KCNQ3* (1); *TRPM3* (1) |
| Ligand gated | *GRIN1* (9); *GRIN2A* (8); *GRIN2B* (8); *GABRG2* (4); *GABRB3* (3); *GABRA3* (2); *GRIN2D* (2); *GABRA1* (1); *GABRB2* (1); *GRIA3* (1) |
| Potassium channel | *KCNQ2* (22); *KCNT1* (11); *KCNA2* (9); *KCNB1* (7); *KCNC1* (5); *KCNA1* (2); *KCNMA1* (1); *KCNN2* (1); *KCNK18* (1); *KCNK4* (1); *KCNC2* (1); *KCND3* (1); *KCNQ3* (1) |
| Sodium channel | *SCN1A* (55); *SCN2A* (32); *SCN8A* (16); *SCN3A* (3) |
| Voltage gated ion | *SCN1A* (55); *SCN2A* (32); *KCNQ2* (22); *CACNA1A* (18); *SCN8A* (16); *KCNA2* (9); *KCNB1* (7); *KCNC1* (5); *CACNA1E* (3); *HCN1* (3); *SCN3A* (3); *KCNA1* (2); *CACNA1C* (1); *CACNA1G* (1); *KCNMA1* (1); *KCNN2* (1); *KCNK18* (1); *KCNK4* (1); *KCNC2* (1); *KCND3* (1); *KCNQ3* (1); *TRPM3* (1) |

**Supplementary Table 3 Primary information on all genes analyzed from EGRP.**

| **Gene** | **Patients** | **Median age diagnosis** | **Median age onset** | **Median patient years** |
| --- | --- | --- | --- | --- |
| *STXBP1* | 78 | 2 | 0.25 | 0.98 |
| *SCN1A* | 55 | 1.19 | 0.42 | 4.49 |
| *SCN2A* | 32 | 2 | 0.25 | 1.66 |
| *KCNQ2* | 22 | 0.2 | 0.01 | 2.26 |
| *CACNA1A* | 18 | 2.5 | 1.67 | 2.28 |
| *SCN8A* | 16 | 1.95 | 0.42 | 2.84 |
| *SLC6A1* | 13 | 5.6 | 2.67 | 4.69 |
| *WDR45* | 13 | 3.9 | 1.5 | 3.41 |
| *KCNT1* | 11 | 2.4 | 0.12 | 1.86 |
| *SYNGAP1* | 11 | 4.1 | 2.5 | 2.52 |
| *PRRT2* | 10 | 0.5 | 0.37 | 3.44 |
| *DEPDC5* | 9 | 7.4 | 1.08 | 6.18 |
| *DNM1* | 9 | 1 | 0.5 | 0.51 |
| *GRIN1* | 9 | 2.4 | 0.44 | 5.08 |
| *IQSEC2* | 9 | 4.5 | 2.46 | 6.93 |
| *KCNA2* | 9 | 7 | 0.45 | 3.04 |
| *PCDH19* | 9 | 2.1 | 0.92 | 3.85 |
| *GRIN2A* | 8 | 6.85 | 3.95 | 2.36 |
| *GRIN2B* | 8 | 2.05 | 0.92 | 3.35 |
| *KCNB1* | 7 | 6.7 | 3.5 | 8.54 |
| *MECP2* | 6 | 3.83 | 4 | 4.31 |
| *NPRL3* | 6 | 5.15 | 2.5 | 3.2 |
| *ATP1A3* | 5 | 3.33 | 0.04 | 2.73 |
| *FRRS1L* | 5 | 2 | 1.58 | 1.08 |
| *GNB1* | 5 | 7.3 | 1.75 | 3.29 |
| *KCNC1* | 5 | 17.6 | 10 | 0.02 |
| *KIF1A* | 5 | 2.1 | 1.42 | 1.55 |
| *NEXMIF* | 5 | 7.7 | 1.08 | 1.55 |
| *PURA* | 5 | 7.6 | 2.26 | 10.97 |
| *SLC2A1* | 5 | 3.3 | 1.2 | 5.13 |
| *SMC1A* | 5 | 13.2 | 0.25 | 1.6 |
| *CDKL5* | 4 | 1.7 | 0.08 | 6.27 |
| *FOXG1* | 4 | 4.85 | 0.5 | 3.61 |
| *GABRG2* | 4 | 1.23 | 0.33 | 0.23 |
| *TBC1D24* | 4 | 0.8 | 0.15 | 2.27 |
| *TBCK* | 4 | 2.4 | 0.71 | 3.57 |
| *TUBA1A* | 4 | 0.75 | 0.3 | 3.4 |
| *16p13.11_del* | 3 | 12 | 10 | 9.16 |
| *ANKRD11* | 3 | 4.5 | 1 | 5.19 |
| *ARID1B* | 3 | 7.3 | 6.17 | 10.81 |
| *ATP1A2* | 3 | 4.5 | 0.78 | 8.82 |
| *CACNA1E* | 3 | 0.8 | 0.5 | 0.75 |
| *CHD2* | 3 | 11.4 | 5.29 | 4.53 |
| *CLTC* | 3 | 11 | 5.5 | 2.08 |
| *GABRB3* | 3 | 2.3 | 0.85 | 3.93 |
| *GLUD1* | 3 | 1.5 | 0.63 | 5.04 |
| *HCN1* | 3 | 2.3 | 2.7 | 3.9 |
| *NF1* | 3 | 1.7 | 3 | 4.84 |
| *PAFAH1B1* | 3 | 0.5 | 0.4 | 2.2 |
| *RHOBTB2* | 3 | 1.6 | 0.17 | 1.29 |
| *SCN3A* | 3 | 11.2 | 0.04 | 1.44 |
| *SPATA5* | 3 | 1.8 | 1.4 | 6.56 |
| *SPTAN1* | 3 | 3 | 0.67 | 0.27 |
| 22q11.2 del | 2 | 0.67 | 1.58 | 5.1 |
| *ABCC8* | 2 | -- | -- | 5.31 |
| *ALG13* | 2 | 5.35 | 0.55 | 6.59 |
| *AP2M1* | 2 | 6.25 | 2.15 | 0.87 |
| *ARFGEF1* | 2 | 14.66 | 3.25 | 13.7 |
| *ATP6V0A1* | 2 | 10.8 | 3 | 9.81 |
| *CASK* | 2 | 1.15 | 1.5 | 0.7 |
| *CSTB* | 2 | 9.95 | 4.29 | 6.04 |
| *CTNNB1* | 2 | 6.55 | -- | 1.61 |
| *DCX* | 2 | 10.9 | 9.5 | 11.17 |
| *DDX3X* | 2 | 5.1 | 3 | 4.34 |
| *DYNC1H1* | 2 | 8 | 0.32 | 11.52 |
| *FGF12* | 2 | 4.28 | 0.02 | 4.99 |
| *GABRA3* | 2 | 23.75 | 0.42 | 4.88 |
| *GRIN2D* | 2 | 10.9 | 0.72 | 8.1 |
| *KANSL1* | 2 | 10.15 | 6.5 | 5 |
| *KCNA1* | 2 | 0.33 | 0.54 | 5.63 |
| *MACF1* | 2 | 21.5 | 4.5 | 15.24 |
| *MYO15A* | 2 | 9.05 | 7 | 7.43 |
| *NBEA* | 2 | 14.85 | 3 | 11.2 |
| *NRXN1* | 2 | 4.5 | 2.25 | 5.1 |
| *NRXN3* | 2 | 3 | 1.17 | 3.57 |
| *PACS1* | 2 | 10.5 | 2.19 | 3.6 |
| *PNKP* | 2 | 1.75 | 0.5 | 4.04 |
| *POGZ* | 2 | 11.45 | 3 | 10.06 |
| *PPP3CA* | 2 | 1.85 | 0.56 | 1.52 |
| *SLC6A5* | 2 | 0.25 | 0 | 3.72 |
| *STX1B* | 2 | 5.4 | 1.71 | 6.54 |
| *SZT2* | 2 | 1.95 | 0.25 | 1.38 |
| *TBR1* | 2 | 12.8 | 11 | 2.19 |
| *TCF4* | 2 | 1.8 | 0.42 | 2.23 |
| *TSC1* | 2 | 3.35 | 1.04 | 0.54 |
| *TUBB2B* | 2 | 5.45 | 0.86 | 6.75 |
| *UBE3A* | 2 | 2.2 | 1.04 | 6.65 |
| *UGDH* | 2 | 0.1 | 0 | 1.34 |
